# Impact of temperature on Covid 19 in India

**DOI:** 10.1101/2020.08.30.20184754

**Authors:** Manas Pratim Roy

**Author notes:** Corresponding address: Dr. Manas Pratim Roy, Ministry of Health and Family Welfare, New Delhi. Source(s) of support: Nil. Conflicting Interest – No. ORCID: 0000-0001-6308-8351.

## Abstract

The role of temperature in Covid 19 pandemic has been subjected to frequent review. An effort was made to find out such relations from three districts in India. Data were analyzed for 14 weeks. It appears that temperature could impact the spread of the pandemic.

## Impact of temperature on Covid 19 in India

The role of temperature in Coronavirus Disease 19 (Covid 19) pandemic has been examined in different parts of the world, including India. (Li et al., 2020; Luo et al., 2020; Roy, 2020) However, such observation for a long period is not common in Indian literature. One multi-centric study discussed the changes between March to May 2020 from six cities in India and found a positive relationship between temperature and Covid 19. (Kumar, 2020) However, the relation during the rainy season is yet to be documented. Therefore, the present analysis tried to find out the impact of temperature on Covid 19, taking account of the rainy season.

For analysis, three districts from different parts of India were considered – Kolkata (east), Bhopal (central), and urban Bengaluru (south). Till 17th August, Kolkata, Bhopal, and Bengaluru recorded more than 32,867 cases; 8462 cases, and 94,106 cases, respectively. Data were retrieved from the website of State Governments. (Govt. of West Bengal, Govt. of Madhya Pradesh, Govt. of Karnataka) The period 5th May – 17th August 2020 was considered. Daily maximum and minimum temperatures were taken into account. (AccuWeather) The difference between the maximum and minimum temperature, termed as Daily Temperature Range (DTR) was also included. These three parameters were compared with daily new positive cases. The correlation coefficient was calculated. Taking a lag period of one week, a similar exercise was done again. Where data were missing, an average of two days was taken for plotting the graphs.

Fig 1 shows a comparison of three districts in terms of the rise in daily new Covid cases. Bengaluru witnessed a disproportionate rise from the last week of June. It seems that the epidemic is experiencing a plateau now in Kolkata and Bhopal.

**Figure 1.**
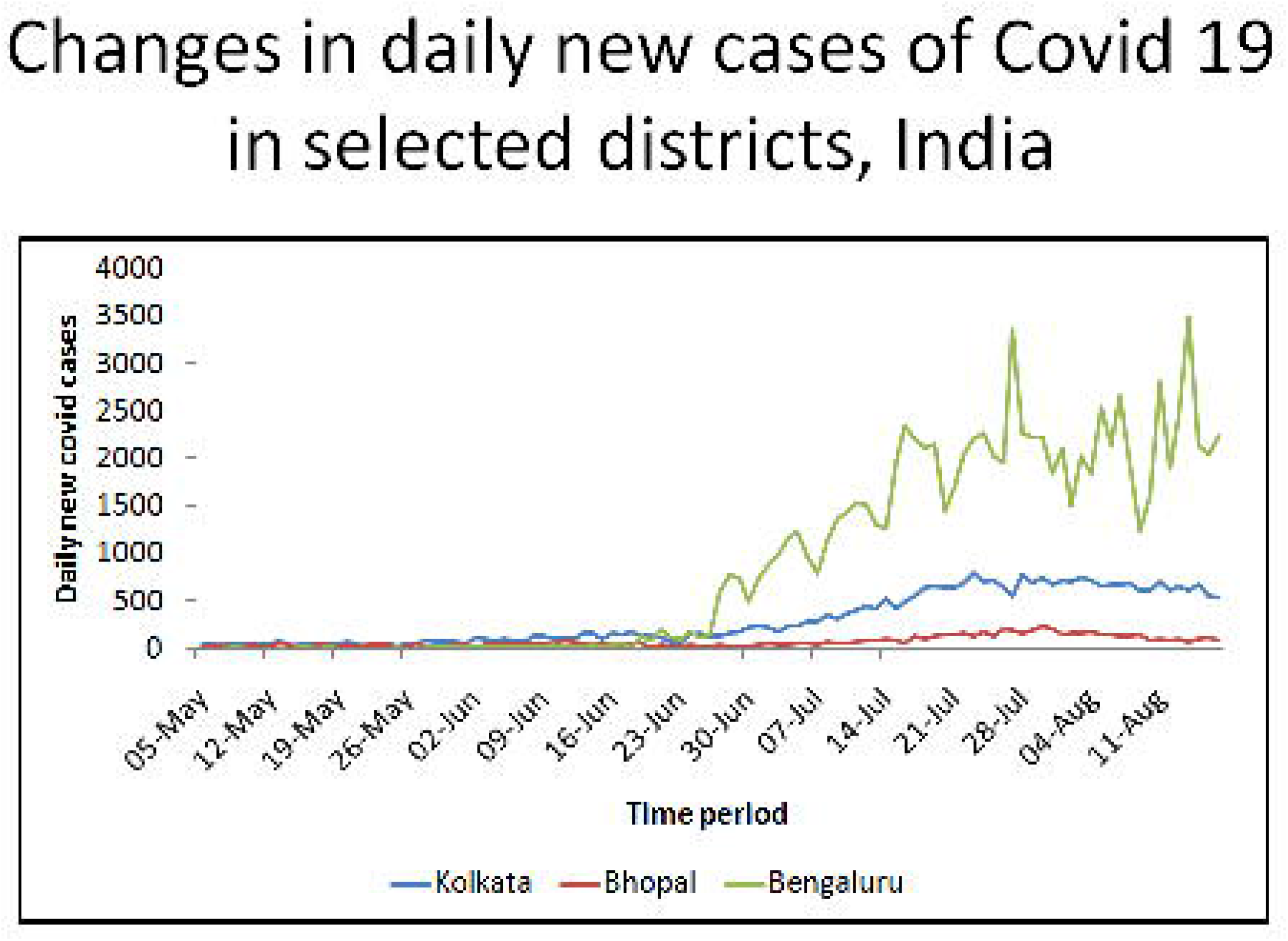

The weekly growth rate was also plotted against the time frame. (Fig 2) Interestingly, it was noted that Bengaluru witnessed sharp peaks and falls before settling around one from mid-July. For Bhopal, the growth rate has been well below one from the 1st part of August. For Kolkata, from a peak of 1.5 at May-end, the growth rate came down by June-end, only to rise again to 1.6 within two weeks, followed by a gradual decline to less than one, suggesting a decline.

**Figure 2.**
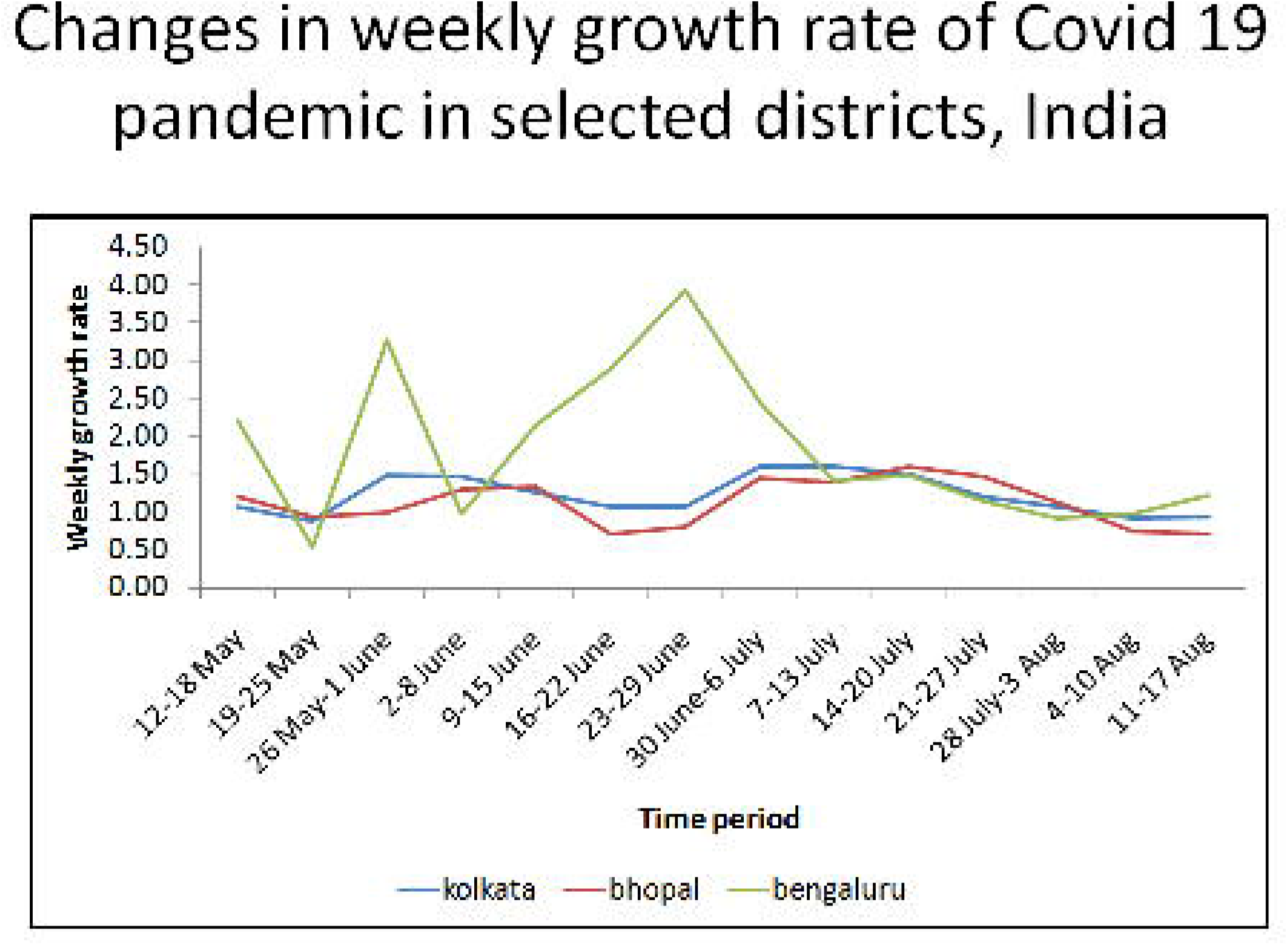

From Table 1-3, it is evident that maximum temperature and DTR share negative relationships with the pandemic, suggesting a rise in any of these two factors will reduce the spread. In the case of Bengaluru, the maximum temperature recorded a strong negative correlation with the number of new Covid cases (r = − 0.765). The extent of impact remained almost similar after taking a lag period of one week. Except for Kolkata, minimum temperature cast a negative influence on the spread of Covid 19.

**Table 1:** Relationship between Covid 19 transmission and temperature, Kolkata

|  | Correlation coefficient with<br>number of daily case | Correlation coefficient with<br>number of daily case (One<br>week lag) |
| --- | --- | --- |
| Maximum Temperature | – 0.198 | – 0.206 |
| Minimum Temperature | 0.300 | 0.253 |
| Daily Temperature Range | – 0.416 | – 0.396 |

**Table 2:**
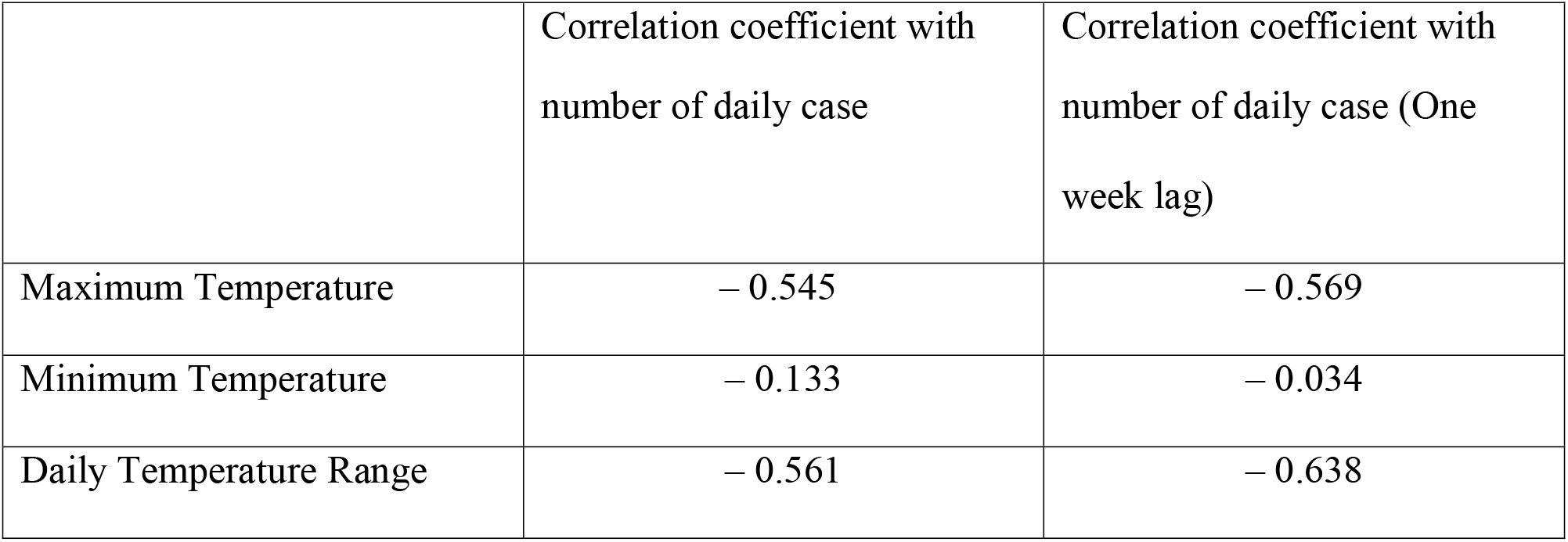
Relationship between Covid 19 transmission and temperature, Bhopal

**Table 3:** Relationship between Covid 19 transmission and temperature, Bengaluru

|  | Correlation coefficient with number of daily case | Correlation coefficient with number of daily case (One week lag) |
| --- | --- | --- |
| Maximum Temperature | – 0.765 | – 0.764 |
| Minimum Temperature | – 0.556 | – 0.547 |
| Daily Temperature Range | – 0.577 | – 0.559 |

The country witnessed lockdown till 31st May 2020. Even after that, the State Governments imposed lockdown from time to time. It could have impacted different patterns of the epidemic in different districts. Another reason between three districts could be explained partially by varying testing rates. While for Karnataka, where Bengaluru is located, it is 36,066/million, it is 15,728/ million for West Bengal, where Kolkata is and 14,088/million for Madhya Pradesh, where Bhopal is located. The higher testing rate could be an explanation for the higher concentration of cases at Bengaluru. (COVID19INDIA) There has been a gradual rise in the number of tests being done per day across the country. It is expected that with time, surveillance has become better. The rise in the number of cases is thus expected with time.

The present study supports the pattern when a reduction in maximum temperature encourages viral transmission but a strong relationship (r > 0.7) could not be found except in Bengaluru. DTR seems to have a moderate to strong influence on the number of daily cases. An analysis from multiple cities in China could not demonstrate any effect of mean temperature on the number of daily cases when the temperature is above 3°C. (Xie et al, 2020) It is assumed that summer and rainy season is not going to have much impact on slowing down the pandemic in India. (Kumar, 2020) There is a need for an in-depth analysis to examine whether any convincing relationship exists between temperature and the present Covid 19 pandemic. Several other factors like migration, containment policy, adherence to lockdown, and personal hygiene may impact the course of the pandemic. All these should be taken into account while conducting future studies on weather and Covid pandemic.

## Data Availability

All data are available in public domain

## References

1. AccuWeather. URL. https://www.accuweather.com/en/in/kolkata/206690/weather-forecast/206690, https://www.accuweather.com/en/in/bengaluru/204108/weather-forecast/204108, https://www.accuweather.com/en/in/bhopal/204408/weather-forecast/204408 [Accessed 28 August, 2020]

2. COVID19INDIA. URL. https://www.covid19india.org/ [Accessed 23 August, 2020]

3. Govt. of Karnataka. COVID 19 information portal: Media Bulletin. URL. http://https://covid19.karnataka.gov.in/govt_bulletin/en/ [Accessed 27 August, 2020].

4. Govt. of Madhya Pradesh. State portal for COVID 19 monitoring. http://mphealthresponse.nhmmp.gov.in/covid/health-bulletin/ [Accessed 27 August, 2020].

5. Govt. of West Bengal. Corona Bulletin. https://www.wbhealth.gov.in/pages/corona/bulletin. [Accessed 27 August, 2020].

6. Kumar S. Effect of meteorological parameters on spread of COVID-19 in India and air quality during lockdown. Sci Total Environ 2020 Nov 25; 745: 141021.

7. Li Q, Guan X, Wu P, et al. 2020. Early Transmission Dynamics in Wuhan, China, of Novel Coronavirus-Infected Pneumonia. N Engl J Med. 382(13):1199-1207. doi:10.1056/NEJMoa2001316

8. Luo W, MS Majumder, D Liu, et al. 2020. The role of absolute humidity on transmission rates of the COVID-19 outbreak. medRxiv. published online February 17. DOI: 10.1101/2020.02.12.20022467

9. Roy MP. 2020. Temperature and Covid 19: India. BMJ Evidence-Based Medicine. http://dx.doi.org/10.1136/bmjebm-2020-111459

10. Xie J, Zhu Y. 2020. Association between ambient temperature and COVID-19 infection in 122 cities from China. Sci Total Environ. 724:138201. doi: 10.1016/j.scitotenv.2020.138201

